# COVID-19 Vaccine’s Gender Paradox

**DOI:** 10.1101/2021.03.26.21254380

**Authors:** Vincenzo Galasso, Paola Profeta, Martial Foucault, Vincent Pons

**Affiliations:** Bocconi University. via Roentgen 1, Milan 20136, Italy; Sciences Po, 98 rue de l’Université, 75007 Paris, France; Harvard Business School, Soldiers Field, Boston MA 02163, United States; Dondena Center; CESIfo; CEPR; IGIER; APE- Baffi Center; Axa Gender Lab; CEVIPOF; CNRS; NBER; Abdul Latif Jameel Poverty Action Lab

## Abstract

Women die less than men of COVID-19, but have been more concerned about its health consequences and more compliant with the public health rules imposed during the pandemic. Since return to normal life depends on vaccination, but delays in acceptance or outright refusals of vaccination are already evident, we investigate gender differences in attitudes and expected behaviors regarding COVID-19 vaccination. Using original data from a survey conducted in December 2020 in ten developed countries (N=13,326), we discover a *COVID-19 Vaccine’s gender paradox*. Being more concerned about COVID-19 and more likely to believe to be infected and consequently to become seriously ill, women could be expected to be more supportive of vaccination than men. Instead, our findings show that women agree less than men to be vaccinated and to make vaccination compulsory. Our evidence suggests that their vaccine hesitance is partly due skepticism, since women are less likely to believe that vaccination is the only solution to COVID-19 and more likely to believe that COVID-19 was created by large corporations. Using a survey experiment performed in these ten countries, we show that information provision on the role of vaccination to become immune to COVID-19 is effective in reducing vaccine hesitance.

## Main Text

The anxious waiting for COVID-19 vaccines has finally come to an end. On December 2nd 2020, UK regulators granted an emergency use authorization for the COVID-19 vaccine tested in a large clinical trial.^1^ Other authorizations by the US FDA (Food & Drug Administration) and the EMA (European Medicines Authority) immediately followed. Starting in September 2020, the rate of transmission of COVID-19 had largely increased in many countries, with natural herd immunity being far from a reality.^2^ Hopes to return to normal life, after almost a year of extraordinary large health, economic and psychological costs,^3,4^ were all placed on the development of a vaccine. The main challenges to curb the pandemic seem now to be the production and storage of a sufficiently large supply of doses of effective vaccines and the launch of an unprecedented vaccination campaign able to reach a large share of the population across the world in the shortest possible time.^5,6^ Since demand will exceed supply for some time, priorities are being established on who should be vaccinated first. However, experience with previous vaccinations shows that countries often struggled with vaccine hesitancy, despite the availability of vaccination services.^7^ Early studies suggest that COVID-19 will be no exception.^8,9,10,11^ Delay in acceptance or outright refusal of vaccination may be due to several reasons, such as complacency towards the disease, a lack in confidence in the vaccine and convenience of the vaccination.^12,13^

So far, the pandemic has been quite gendered. More men than women are dying of COVID-19 and attitudes and behaviors related to COVID-19 differ also across gender.^14,15^ Women have been more concerned about the health consequences of COVID-19 and more compliant with the public health rules imposed during the pandemic.^16^ In this study, we investigate gender differences in attitudes and expected behaviors regarding COVID-19 vaccination. We use original data from a nationally representative survey conducted in ten developed countries to analyze gender differences in the belief that vaccination is the only permanent solution to the pandemic, as well as in the agreement with being vaccinated and with compulsory vaccination.

### Survey Data

Our survey data^17^ cover ten countries, Australia (N = 1,006), Austria (N = 994), France (N = 2,121), Germany (N = 2,091), Italy (N = 1,025), New Zealand (N = 1,011), Poland (N = 1,023), Sweden (N = 1,016), the United Kingdom (N = 1,031), and the United States (N = 2,008), for a total of 13,326 respondents. All these countries have high income per capita and advanced health systems, allowing us to pool their data in a common analysis. However, the pandemic affected them very differently, which increases the external validity of our findings. By the end of 2020, Italy, the United Kingdom and the United States were among the countries with the higher mortality per 100 thousands people,^18^ whereas New Zealand and Australia were among the countries with the lower rates (see Table S1). The survey was administered between December 2^nd^ and December 10^th^ 2020, when most countries were experiencing the second wave of the pandemic and new lockdown measures – particularly targeted for the Holidays season, were imposed. Meanwhile, vaccination plans were underway in most countries.

The survey collects information on individuals’ attitudes and behavior towards COVID-19 per se and towards COVID-19 vaccination. More specifically, respondents were asked how serious they believe the health consequences of COVID-19 to be in their country. They were asked to report their current level of compliance with several COVID-19 related health and social distancing rules. Additional questions concerned COVID-19 vaccination. The survey collected also a wide range of sociodemographic and attitudinal factors.

### Gender Differences in Compliance

Using December 2020 data from all ten countries (N=13,321), we observe large gender differences in the individual perceptions regarding the seriousness of COVID-19 as a health problem in the respondent’s country (M = 0.317 vs. 0.273, Mdiff = 0.044, 95% CI [0.028; 0.059]). Fig. S1 displays the results by country, separately for men and women. The large differences across countries reflect differences in the actual magnitude of the pandemic (see Table S1).

Important gender differences emerge also in the compliance with public health rules. Individuals were asked to evaluate how strictly they were following nine recommended rules on a 0–10 scale (from “not at all” to “completely”): washing hands more often, coughing into one’s elbow, ending the greeting of people by shaking hands or hugging, avoiding crowed places, keeping physical distance from others, staying at home, stopping visits to friends, wearing face masks in public places and leaving home less than once a day. We construct an overall index of respondents’ compliance with public health rules. Pooling data from all ten countries (N=13,326), compliance to the rules was markedly larger among women than among men (M = 0.759 vs. 0. 710, Mdiff = 0.049, 95% CI [0.041; 0.056]). Figure 1 displays our compliance index separately for men and women by country using December 2020 data. Gender differences are apparent (and statistically significant) in almost all countries. Interestingly, when compared to previous studies,^16^ the compliance index is rather stable over time, from 0.747 in April 2020 to 0.735 in December 2020.

**Fig. 1.**
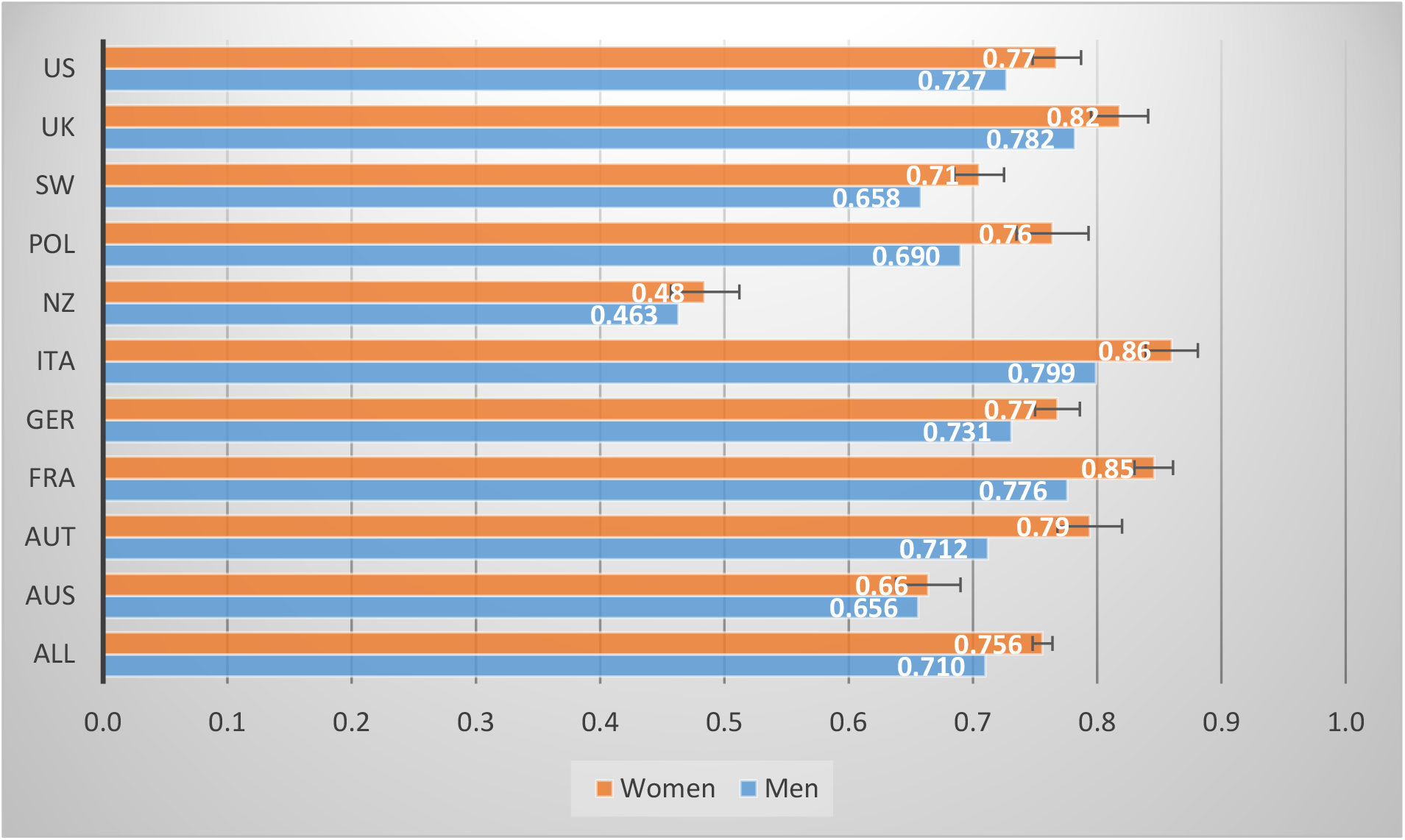
Compliance Index. Compliance Index for men and women, in the pooled sample and by country. 95% confidence intervals from regressions of the Compliance Index on female are reported.

### Gender Differences in Vaccination

Does women’s more protective behavior during the pandemic induce them to agree more with COVID-19 vaccination? Our survey allows us to learn about individuals’ beliefs, attitudes and intentions towards COVID-19 vaccination. Respondents were asked how much they believe that the only permanent solution to this pandemic is developing a vaccine, on a 0-10 scale (from completely unlikely to very likely). For comparability with previous measures, we normalize the index between 0 and 1. Substantial gender differences emerge in individual attitudes towards the vaccine. Pooling data from all ten countries (N=13,019), we find that women are much less likely to believe the vaccine to represent the only permanent solution to the pandemic (M = 0.690 vs. 0.7271, Mdiff =-0.036, 95% CI [-0.047; −0.026]). Fig. S2 displays these individual attitudes by country, separately for men and women.

To investigate possible gender differences in vaccination behavior, we use a question in our survey, in which respondents were asked whether they would agree to be vaccinated, if a vaccine against COVID19 was available in the next few months, on a 0-10 scale (from not at all likely to extremely likely). Again, we normalize the index between 0 and 1. Our results show a large and significant gender difference in the willingness to be vaccinated. Pooling data from all ten countries (N=11,918), we show that women are much less likely to agree to be vaccinated (M = 0.581 vs. 0.664, Mdiff = −0.083, 95% CI [-0.095; −0.071]). Figure 2 displays this vaccination agreement index by country, separately for men and women. Gender differences are large and significant in every country.

**Fig. 2:**
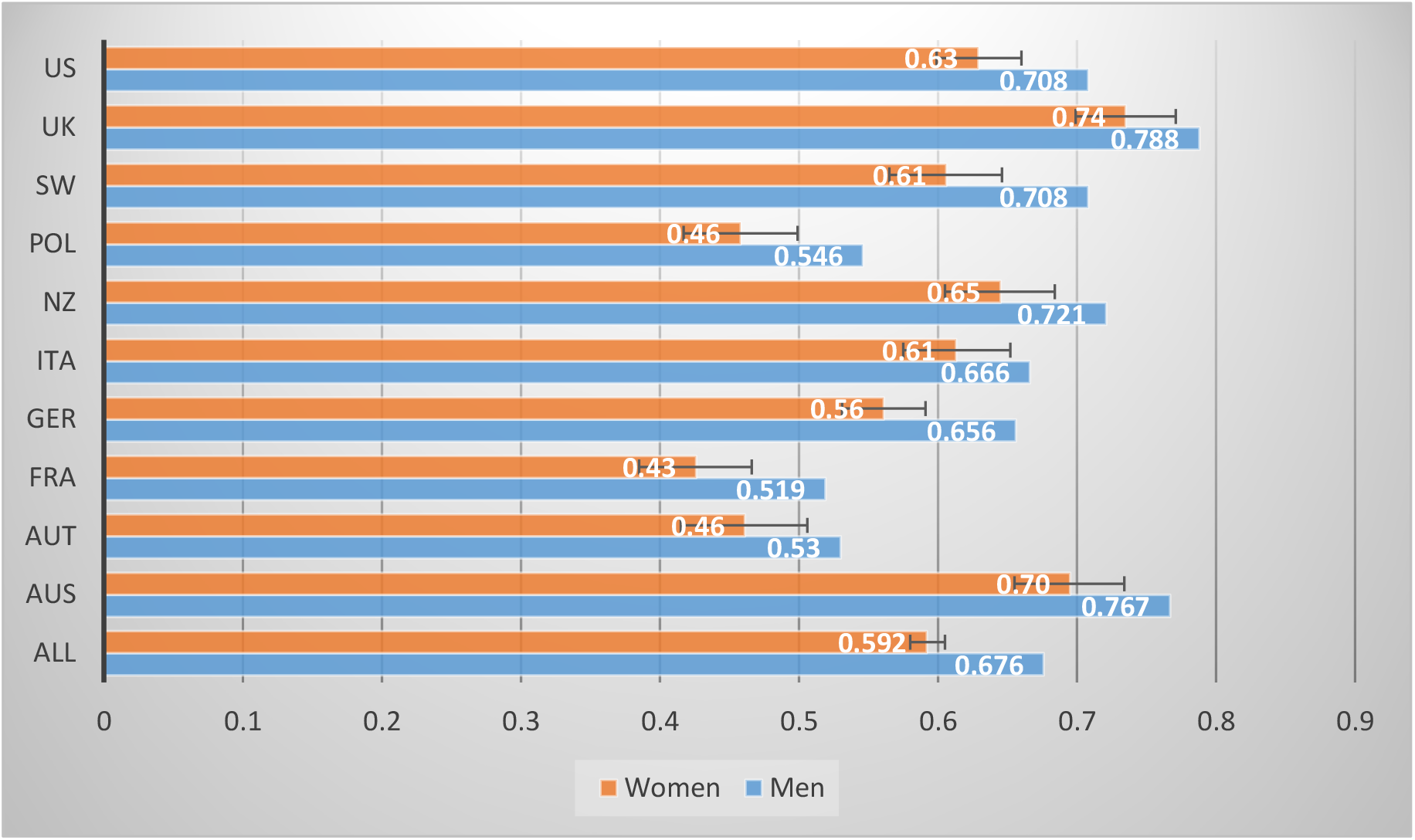
Vaccination Agreement Index. Vaccination Agreement Index for men and women, in the pooled sample and by country. 95% confidence intervals from regressions of the Vaccination Agreement Index on female are reported.

With the vaccination agreement index ranging from only 0.470 in France to 0.761 in the UK, policy-makers may consider to impose compulsory vaccination. We use a question in our survey to evaluate the support for such a policy. Respondents were asked if they agree that being vaccinated should be compulsory, since public health reasons are more important than respect for individual freedom of choice, or viceversa. Pooling data from all ten countries (N=12,126), we show that women are much less likely to agree with compulsory vaccination (M = 0.377 vs. 0.443, Mdiff = −0.066, 95% CI [-0.083; −0.048]). Fig. S3 shows these results by country and gender.

### COVID-19 Vaccine’s Gender Paradox: Explanatory Factors

Our results indicate the existence of a COVID-19 vaccine’s gender paradox: women are more concerned about COVID-19, more compliant with the related public health rules, but less willing to get the vaccine. What are the determinants of these gender differences with respect to COVID-19 vaccination? Vaccine hesitancy is a complex matter. The WHO–SAGE vaccine hesitancy working group suggested that complacency towards the disease, a lack in confidence in the vaccine, and vaccination convenience are important factors that may hinder vaccination.^12,13^ Complacency refers to the risks of vaccine preventable diseases being perceived as low, so that vaccination is not deemed necessary. Convenience relates to physical availability, affordability and appeal of the immunization service. Confidence requires trust in the effectiveness and safety of the vaccine,^19^ in the health services delivering it and in the motivations of the policy-makers launching the vaccination campaign.^20^

Gender differences in sociodemographic characteristics or employment status may create different perceptions about these components and induce different behaviors. For instance, women may be more concerned about COVID-19, more compliant with the rules and less complacent towards COVID-19 if they are older or if they perform an economic activity that expose them to a higher risk of contagion.^21,22^ To account for these confounding factors, we include a set of control variables in our regression analysis. Our outcome variables of interest are: (i) the individuals’ beliefs that the vaccine represents a permanent solution to the pandemic; (ii) their agreement to be vaccinated; (iii) their agreement to compulsory vaccination. We regress each of these three variables on the female dummy and on a set of control variables: dummy variables for age groups, level of education, type of occupation (blue collar, service, white collar, no occupation) and geographical locations (specifically, country-region fixed effects).

Figure 3 plots estimates of gender differences in our pooled sample for the three main outcomes of interest, with no individual controls (only country-region fixed effects) and after controlling for the sociodemographic variables too. We report the exact point estimates for the pooled sample in Table S2. Even after controlling for a large number of sociodemographic characteristics, women remain much less likely than men to believe that vaccination is the solution to COVID-19. They agree less with being vaccinated and with making vaccination mandatory. Tables S3 to S5 report the results of our empirical analysis on the three outcome variables separately for each country.

**Fig. 3.**
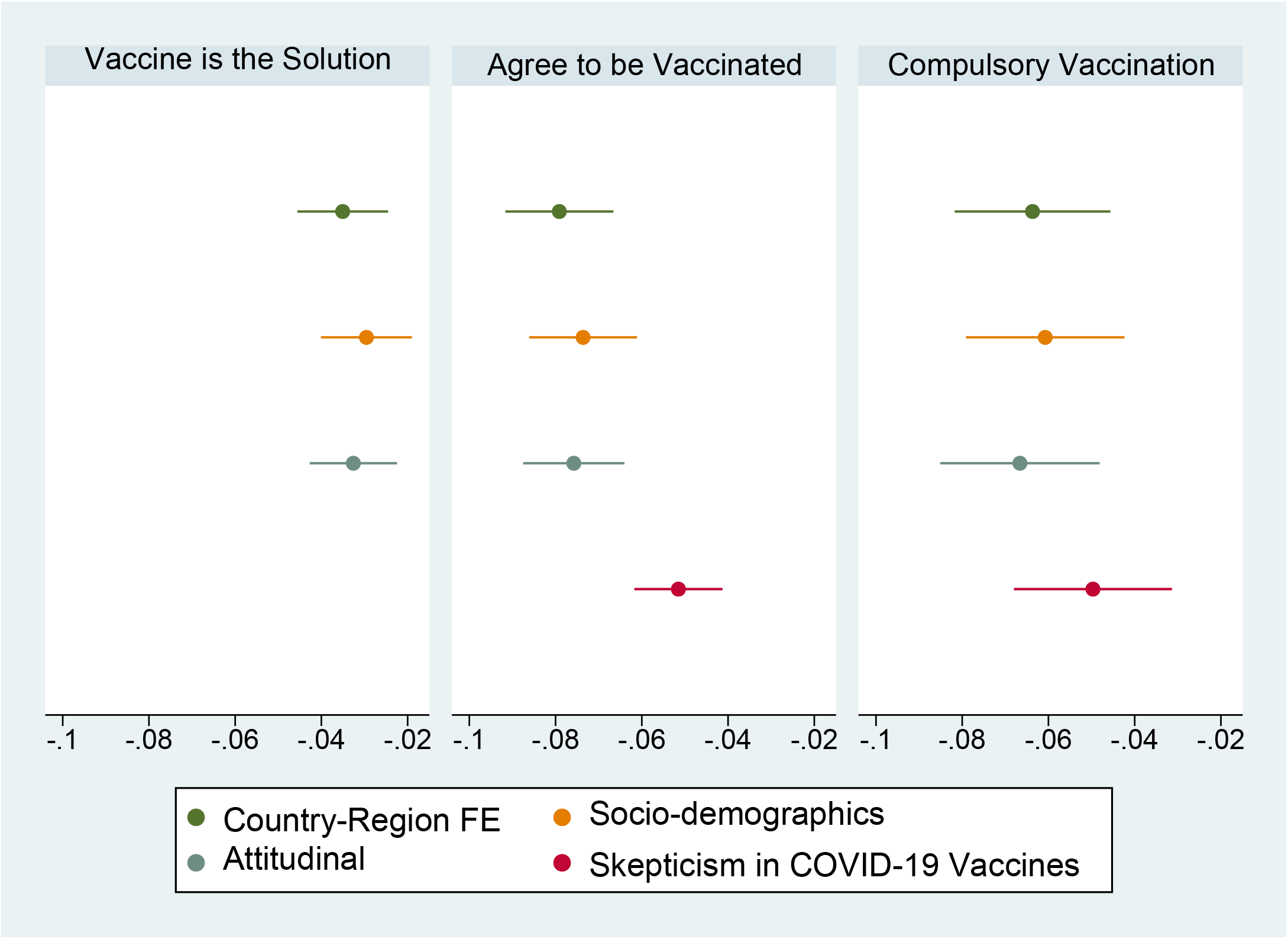
Gender Effects in Vaccination Outcomes. Point estimates of the female coefficient, and 95% confidence intervals, from regressions using pooled data of respectively the variables “Vaccine is the Solution”, “Agree to be Vaccinated” and “Agree to Compulsory Vaccination” on the female coefficient and on Country-Region FE and then adding Sociodemographic factors, Attitudinal factors and Skepticism in COVID-19 vaccines.

We then investigate whether this COVID-19 vaccine’s gender paradox depends on gender differences in attitudinal factors.^23^ We consider four factors affecting individual complacency:^12,13^ the perceived probability of becoming infected, the perceived probability of becoming seriously ill if infected, the assessment of the seriousness of COVID-19 as a health problem and the level of risk aversion. We analyze also two factors related to the confidence in the vaccine: trust towards scientists, who are ultimately responsible for effectiveness and safety of the vaccines,^19^ and political ideology, which measures support for government intervention and affects the degree of alignment with the existing government. The latter element may capture the confidence in the motivations of the policy-makers.^20^

To study gender differences in these six attitudinal factors, we regress each indicator on the female dummy, on the country-region fixed effects and on the socio-demographic controls. The results, reported in Table S6, show that women are more concerned with the health consequences of COVID-19, more likely to believe to be infected and to become seriously ill, if infected. Moreover, they are more risk averse than men. In line with the existing literature,^16,24^ these findings suggest that women are certainly less complacent than men about COVID-19 health risks. Women are also more left-leaning than men – in accordance with the political science literature.^25,26^ No gender differences emerge in the trust towards scientists.

The results of augmented regressions controlling for these six additional factors, shown in Figure 3 and in Table S2 (columns 3, 6 and 9), show their importance in explaining the variation in beliefs, attitudes and expected behavior towards COVID-19 vaccination. The R-squared increases substantially, if compared with the regressions that do not include them (see respectively columns 2, 5 and 8 in Table S2). However, the point estimates of the female dummy are not affected by the introduction of these attitudinal elements. If anything, these estimates tend to increase, since women’s individual characteristics, such as their stronger concern about the health consequences of COVID-19, should make them more favorable than men to vaccination.

### Skepticism in COVID-19 vaccine

Is COVID-19 vaccine’s gender paradox due to women being more skeptical about the vaccine? Women are less likely to consider vaccination as the only permanent solution to the pandemic, which may be due to skepticism about its effectiveness or to the belief in the existence of other solutions, such as strict compliance with public health rules to avoid contagion – hence, self-reliance. Skepticism about vaccination may also come from individuals mistrusting the motivation of the policy-makers about the vaccination.^20^ We can capture this aspect using a question in the survey in which respondents were asked how much they believe that the virus has been created by large corporations because some of them can directly profit from it, on a 0-10 scale (from completely unlikely to very likely). Another element that may amplify individuals’ skepticism is their being parents and feeling more responsible about vaccination, as their decision is relevant for their children too. The effect of the parental role on vaccine hesitance seems to be stronger among women.^27^

To measure the relevance of the skepticism in the COVID-19 vaccines, we thus augment our previous regressions with the following three variables: vaccination as the only solution to the pandemic, virus created by large corporations and school age children. Results of these augmented regressions, reported in Figure 3 and in Table S7, show the importance of these variables in explaining the agreement to be vaccinated and to compulsory vaccination. The R-squared largely increase, if compared with the regressions that did not include these factors (see respectively columns 6 and 9 in Table S2). In particular, individuals who believe vaccination to be the only permanent solution to COVID-19 are much more willing to be vaccinated and to agree with compulsory vaccination, whereas those who believe COVID-19 to be created by large corporations are less likely to agree with vaccination. Figure 3 shows that the inclusion of these variables reduces the point estimate of the female dummy, which however remains significant and large. Women are less likely to believe that vaccination is the only permanent solution to COVID-19 and more likely to believe that COVID-19 was created by large corporations. Hence, they agree less than men to be vaccinated.

### Policy Implication

Despite the successful development of COVID-19 vaccines, vaccination hesitancy may delay the eradication of the COVID-19 pandemic. In France, only 28.4% of the respondents answered 7 to 10 to the question on the agreement to be vaccinated. Among the ten countries in our study, only in Australia and in the UK, this percentage reaches 60%. Effective public persuasion campaigns in favor of vaccination are thus strongly needed,^28^ especially to convince women.

We assess the effectiveness of different information campaigns, exploiting a survey experiment embedded in our survey, in which respondents in each country were randomly assigned to five different groups. All respondents were asked “If a vaccine against COVID19 was available in the next few months, would you agree to be vaccinated?” with answers on a 0-10 scale (from not at all likely to extremely likely). Individuals in the control group were asked this question only.

Individuals in every treatment group received the information statement: “The only way to become immune to COVID-19 in the long run is by vaccination”. Moreover, they were primed with different statements on the positive consequences of being vaccinated, respectively for: not being infected (group i), not infecting others (group ii), protecting health in the country (group iii) and protecting the economy (group iv).

Table 1 shows the results of our empirical analysis, in which we regress agreement to be vaccinated on the treatments, on the female dummy, on socio-demographic characteristics and country-region fixed effects. The informational treatment has a positive and statistically significant effect on vaccination agreement (see column 1, which compares all treatment groups to the control group). Receiving the informational message on the role of vaccination increases the agreement of 0.031 point over a sample mean of 0.627. This effect seems stronger for men (0.042) than for women (0.024), as shown in Table 1 (columns 2 and 3), although this difference is not statistically significant. When considering the effect of the different statements on the positive consequences of vaccination, results in Table 1 (column 4) suggests that they are all equally effective. Again, they seem to be more effective for men than for women (see columns 5 and 6), although the difference is not statistically significant.

**Table 1:**
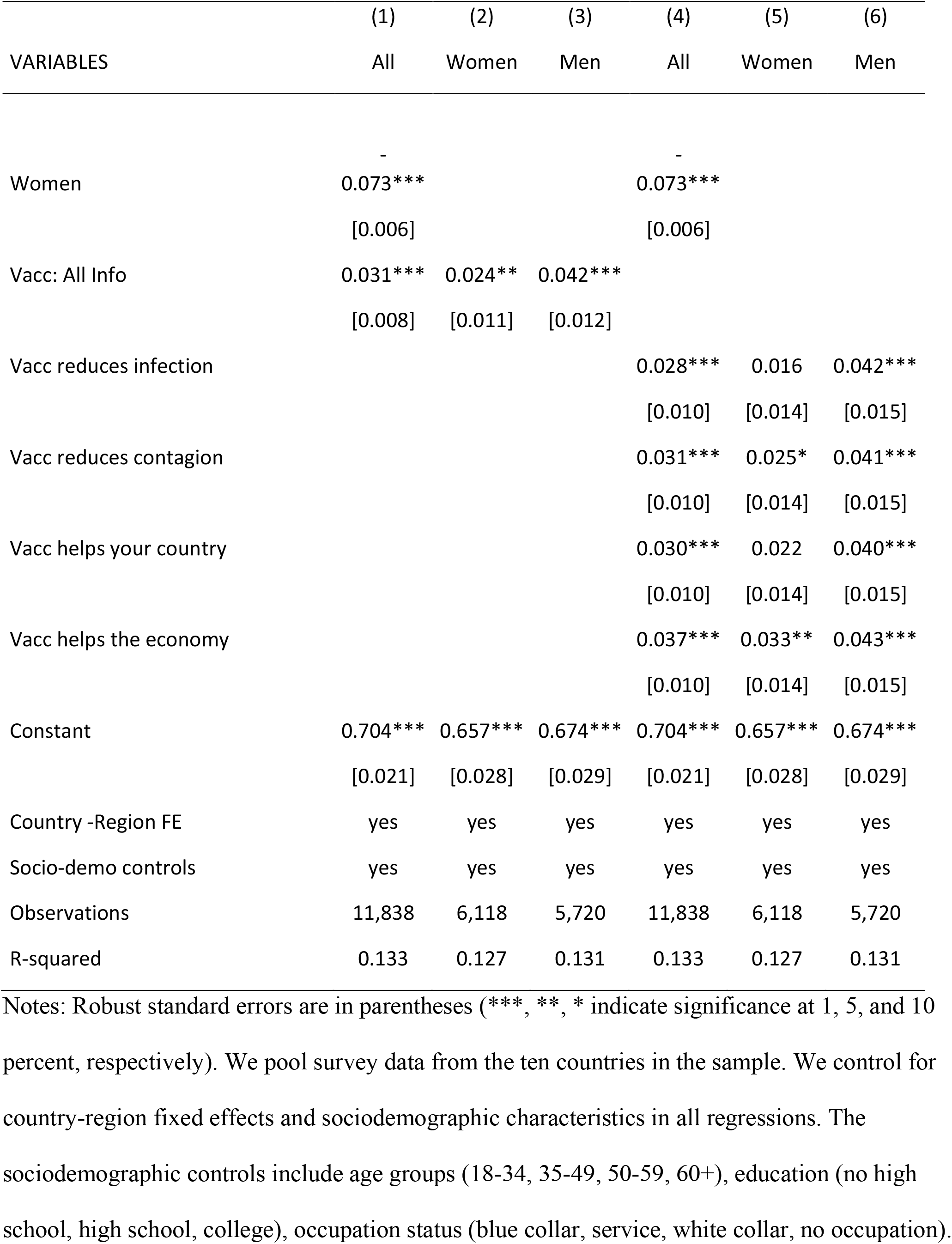
Agreement to Vaccination.

Hence, our findings show that public persuasion campaigns informing people about the unique role of vaccines to immunize against COVID-19 are effective in reducing vaccine hesitancy, yet more so for men. Women-targeted persuasion campaigns may thus be needed.

## Data Availability

Our survey data are not public for now but the data and codes will be made available on the journal webpage upon publication and Galasso′ Harvard dataverse

## Materials and Methods

### Survey Data

We use data from the December 2020 wave of a survey launched contemporaneously in nine countries: Australia, France, Germany, Italy, New Zealand, Poland, Sweden, UK and USA by professional survey companies on representative samples of each country’s citizens. This survey is part of the REPEAT project (REpresentations, PErceptions and ATtitudes on the COVID-19), which collects information on perceptions and individual behavior related to COVID-19 and to the public health measures discussed (or adopted) to limit the diffusion of the virus. Table S1 reports, for each country, the days in which the wave was fielded, the number of observations and number of deaths per 100 thousands people on December 30th 2020.

The outcome variable used in Figure S1 is obtained from the answers to the question “Would you say that the consequences of the coronavirus epidemic for health in your country are today” on a 1-5 scale from “very serious” to “not serious at all”. We created a dummy variable for “very serious”. The outcome variable used in Figure 1 is an index of compliance with some public health measures suggested or applied in the different countries. For each country, respondents were asked nine questions on their level of compliance (on a 0-10 scale, from “not at all” to “completely”) with the following items: (i) washing your hands more often and/or for a longer amount of time; (ii) coughing or sneezing into your elbow or a tissue; (iii) have stopped greeting others by shaking hands, hugging or kissing; (iv) keep a distance of three six feet between yourself and other people outside your home; (v) have reduced your trips outside; (vi) avoid busy places (public transportation, restaurants, sports …); (vii) have stopped seeing friends; (viii) wearing a mask or protection over your nose and mouth when you are outside your home; (ix) leave your home less than once a day on average.. We created a compliance index by averaging these answers for each respondent and normalizing the average on the 0-1 scale.

The outcome variable used in Figure S2 and Tables S2-S3 comes from answers to the question of how much respondents believe, on a 0-10 scale (from completely unlikely to very likely), the following statement to be true: “The only permanent solution to this pandemic is developing a vaccine”. We normalize the index between 0 and 1.

The outcome variable used in Figures 2 and 3 and Tables 1, S2, S4 and S7 is a Vaccination Agreement index constructed using answers to the question “If a vaccine against COVID19 was available in the next few months, would you agree to be vaccinated?” on a 0-10 scale (from not at all likely to very likely). We normalize the index between 0 and 1. In every country, respondents to this question were divided in five groups that received the following formulation of the question:

- Group 1: The only way to become immune to covid-19 in the long run is by vaccination. In this case, if you were vaccinated, you could avoid getting infected with the virus. If a vaccine against COVID19 was available in the next few months, would you agree to be vaccinated?
- Group 2: The only way to become immune to covid-19 in the long run is by vaccination. In this case, if you were vaccinated, you might be able to avoid passing the virus on to others. If a vaccine against COVID19 was available in the next few months, would you agree to be vaccinated?
- Group 3: The only way to become immune to covid-19 in the long run is by vaccination. In this case, if a person was vaccinated, they could avoid getting infected with the virus. This would protect the health of people in (your country). If a vaccine against COVID19 was available in the next few months, would you agree to be vaccinated?
- Group 4: The only way to become immune to covid-19 in the long run is by vaccination. In this case, if a person was vaccinated, they could avoid getting infected with the virus. It would allow a return to normal economic activity in Italy and reduce unemployment. If a vaccine against COVID19 was available in the next few months, would you agree to be vaccinated?
- Group 5: If a vaccine against COVID19 was available in the next few months, would you agree to be vaccinated?

The outcome variable used in Figures 3 and S3 and Tables S2, S5 and S7 is obtained from the following question: “Which of the following two statements do you agree with? (1) Being vaccinated should not be compulsory: respect for individual freedom of choice is more important than public health reasons. (2) Being vaccinated should be compulsory: public health reasons are more important than respect for individual freedom of choice. We create the dummy variable Agreement to Compulsory Vaccination, which takes value 1 for the latter answer and value 0 for the former.

The survey also collected socio-economic and demographic information, such as gender, age, education, geographical location and occupation status. Besides gender, which represents our main variable of interest, we use this information to construct the following variables, which are used as controls in our regressions: age groups (18-34, 35-49, 50-59, 60+), education (no high school, high school, college), occupation status (blue collar, service, white collar, no occupation), macro-regions (by country) and countries.

We use additional information on attitudinal factors and on vaccination related issues. More specifically, we use answers to the question on how easy or difficult is it for you to accept taking risks in health matters (on a 0-10 scale, with 0 being “very difficult” to 10 “very easy”) to construct a measure of risk aversion. We use the question on whether individuals trust scientist (Yes or No) to create a dummy variable for trust in scientists. We use a question “what is the likelihood that you will be infected by COVID19 if you resume your usual way of life (work, leisure, etc.)?” (on 0-10 scale from “very unlikely” to “very likely”) to construct a measure of the subjective probability of being infected. We use a question “In your opinion, what is the likelihood that you would be seriously ill if you were infected by COVID19?” (on 0-10 scale from “very unlikely” to “very likely”) to construct a measure of the subjective probability of being seriously ill, if infected. We use a question on the individual political ideology (on a scale from 0 to 10, where 0 is left and 10 is right) to construct three dummy variables for liberal (0-3), centrist (4-6) and conservative (7-10).

We use answers to the question “In your opinion, how likely is it that the following statement is true: The virus has been created by large corporations because some of them can directly profit from it” on a 0-10 scale (from not at all likely to very likely), to create a variable on multinationals’ fault for COVID-19. Finally, we create a variable for parents having children in schooling age, by using answers to the question “If you have a child in schooling age (attending either primary or secondary education), what schooling level is he/she attending? If you have multiple children in schooling age please refer to the youngest one?” with possible answers being (1) I do not have a child in schooling age; (2) Primary school; (3) Lower Secondary; (4) Upper Secondary. We construct a dummy variable schooling age that takes value 1 for anwers (2) to (4) and value 0 otherwise.

### Methods

To measure the existence of a gender gap in our outcomes of interest (belief that vaccination is a solution to COVID-19; agreement to be vaccinated; agreement to compulsory vaccination), we use OLS estimates of the following linear equation:

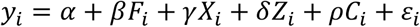

where y_i_ is one of the outcomes of interest, F_i_ is a dummy for female, X_i_ is the vector of control variables that capture the individual sociodemographic factors (age groups, education, occupation status, country-region fixed effects), Z_i_ is the vector of attitudinal characteristics (risk aversion, trust in science, probability of being infected, probability of being seriously ill if infected, political ideology), C_i_ is the vector of variables capturing confidence in COVID-19 vaccines (belief that vaccination is a solution to COVID-19, COVID-19 multinationals’ fault, parents of children in schooling age) and ε is a random error. We use survey weights that ensure the survey to be representative of the population.

These specifications (with or without the controls, X_i_, Z_i_ and C_i_) are used in Tables S1 to S7. Table S2 provides the estimates of the above linear equation on the pooled sample for the three main outcomes (belief that vaccination is a solution to COVID-19; agreement to be vaccinated; agreement to compulsory vaccination). For each outcome, the first column presents the estimate of the equation only with the female dummy and country-region fixed effects, in the second column we add the sociodemographic controls and in the third column also the attitudinal controls.

Tables S3 to S5 provide, one for each of the three main outcomes, the estimates of the above linear equation, with the female dummy, region fixed effects and sociodemographic controls, separately for the pooled sample and for each country. Table S6 provides the estimates of the above linear equation, in which the outcome variables are the different attitudinal factors that are regressed on the female dummy, the country-region fixed effects and the sociodemographic controls. Table S7 provides the estimates of the above linear equation, in which the outcome variables are the agreement to be vaccinated and the agreement to compulsory vaccination, which are regressed on the female dummy, on the country-region fixed effects, on the sociodemographic controls, on the attitudinal controls and also on the variables capturing confidence in COVID-19 vaccines.

To study the informational effect regarding COVID-19 vaccination, we run the following linear equation:

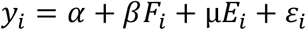

where y_i_ is the variable agreement to be vaccinated, E_i_ is the vector that captures the four different informational treatments, F_i_ is a dummy for female, X_i_ is the vector of control variables that capture the individual sociodemographic factors (age groups, education, occupation status, country-region fixed effects), and ε the error term. Results of this specification are reported in Table 1.

## Acknowledgments

Survey data from the project Attitudes on COVID-19: A Comparative Study, chaired by Sylvain Brouard and Martial Foucault (Sciences Po). Financial Support from ANR (French Agency for Research) - REPEAT grant (Special COVID-19) and Unicredit Foundation are acknowledged. We have complied with all relevant ethical regulations. In every country and each wave of the survey, informed consent was obtained from the respondents by the survey companies IPSOS and CSA.

## Author contributions

Vincenzo Galasso and Paola Profeta did the conceptualization of the research question, the data curation, the formal analysis, and the writing of the paper; Martial Foucault and Vincent Pons provided comments on the final draft. Martial Foucault designed the survey questionnaire; Vincenzo Galasso and Vincent Pons provided comments on the final draft of the questionnaire.

## Data and materials availability

The data description is in the paper. All data, code, and materials used in the analysis will be made available to the reviewers at any time, and to the general public upon publication.

## Supplementary Information

is available for this paper

Correspondence and requests for materials should be addressed to Vincenzo Galasso, Department of Social and Political Sciences, Bocconi University, via Roentgen 1, Milan 20136, Italy

