## Supplementary Material for "COVID-19 Vaccine’s Gender Paradox"

**Affiliations:**

[4] Dondena Center

[5] CESIfo.

[6] CEPR.

[7] IGIER.

[8] APE- Baffi Center.

[9] Axa Gender Lab.

[10] CEVIPOF.

[11] CNRS

[12] NBER

[13] Abdul Latif Jameel Poverty Action Lab

* These authors contributed equally to this work.

This PDF file includes:

Figures S1 and S3

Tables S1 to S7

**Fig. S1**. COVID19 is a very serious health concern

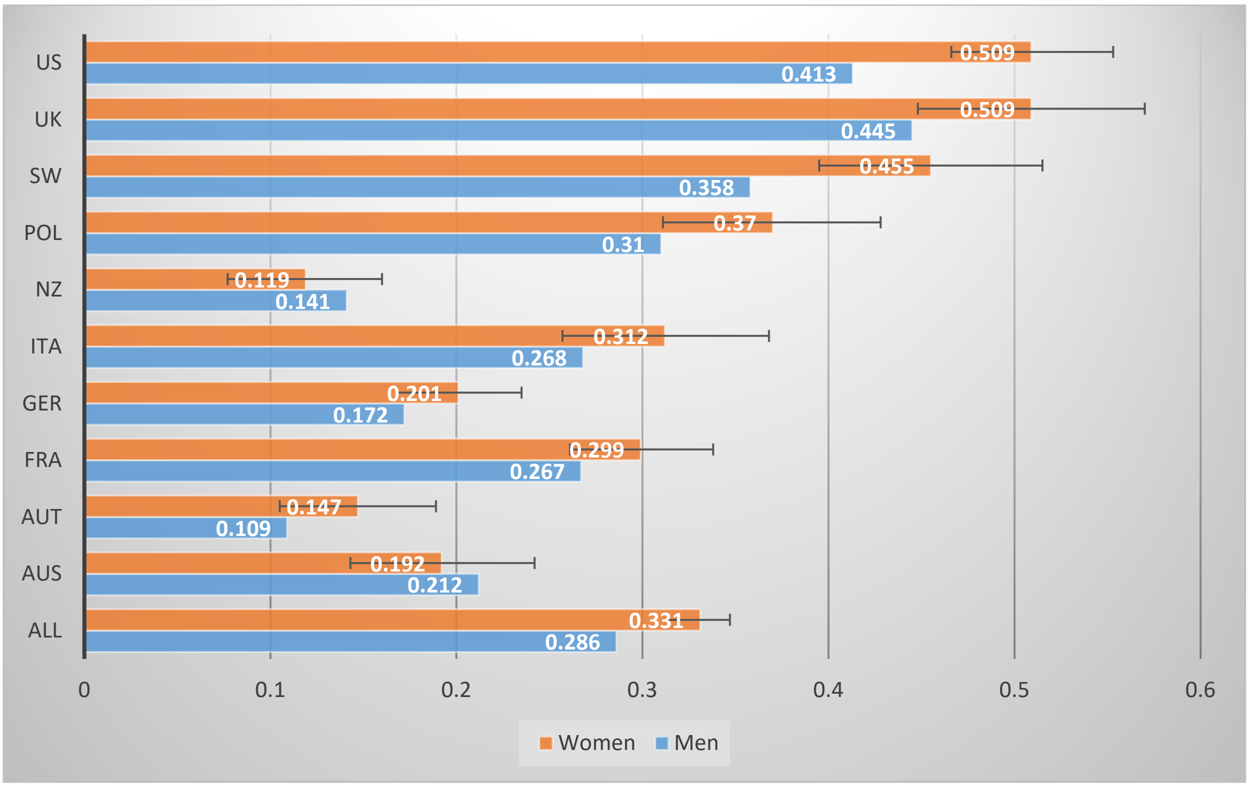

**Fig. S2.** Vaccination is the only Permanent Solution to COVID-19

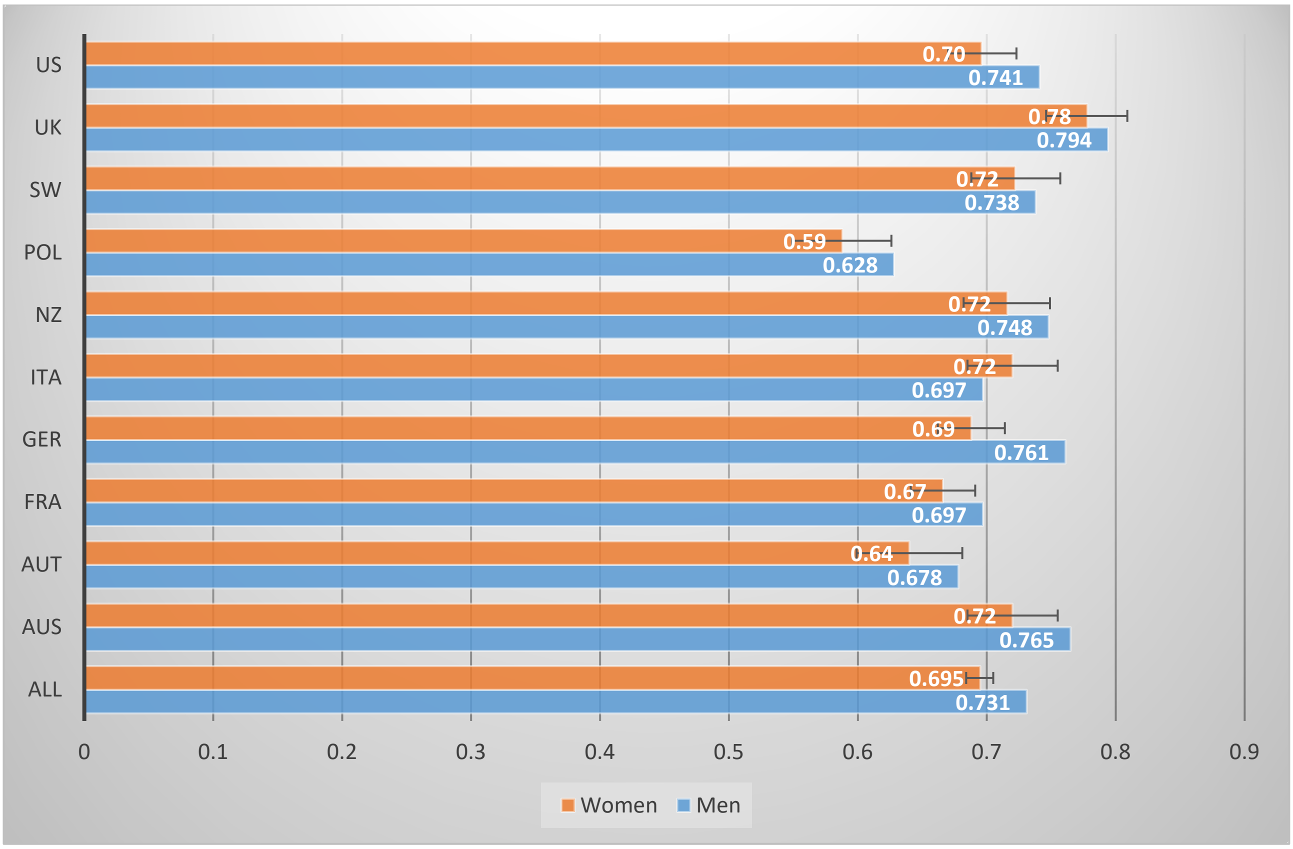

**Fig. S3**. Agreement with Compulsory Vaccination

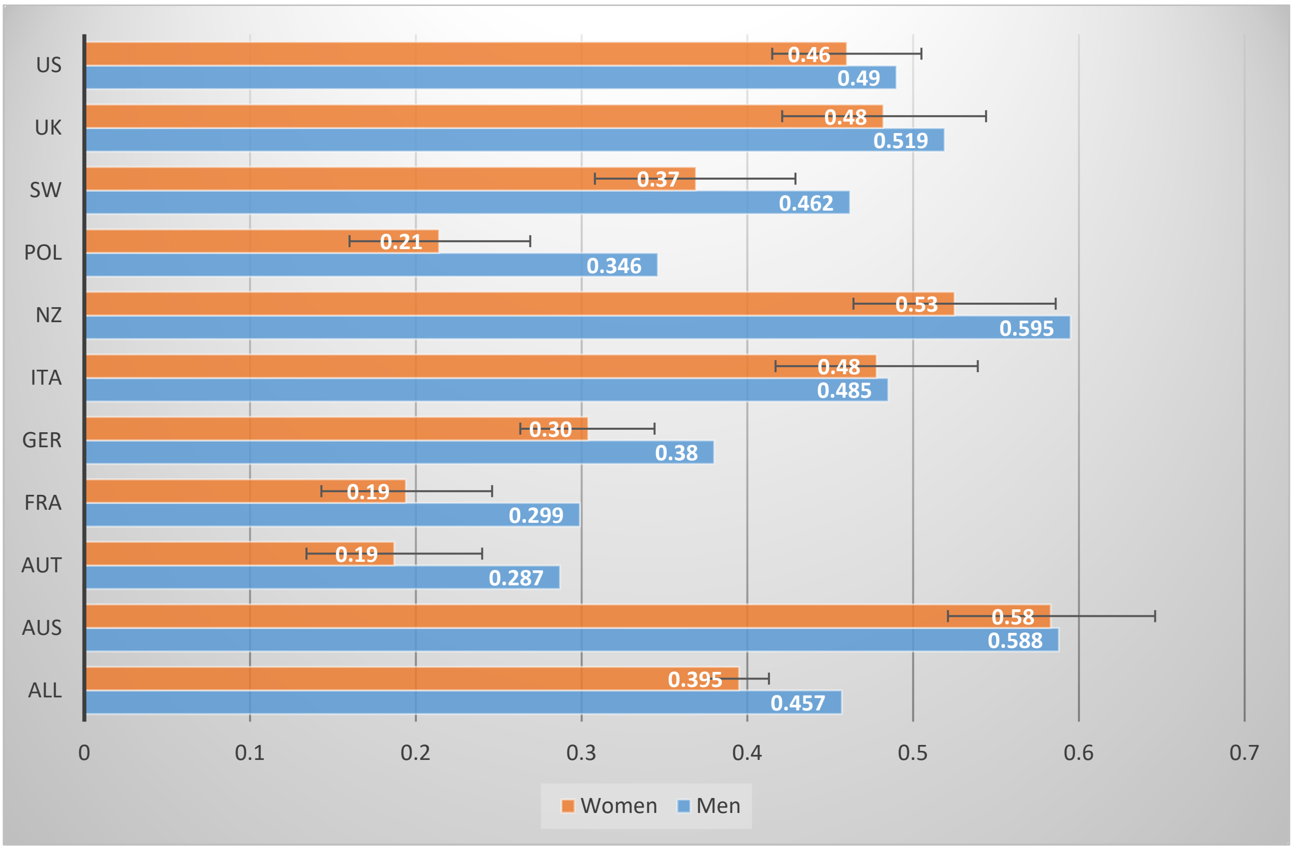

**Table S1.** Survey Dates and COVID-19 Mortality

| | **Dates of the Survey** | **Observations** | **Deaths per 100,000 population** |
|---|---|---|---|
| Australia | 4-10 December 2020 | 1,006 | 3.6 |
| Austria | 5-9 December 2020 | 994 | 68.5 |
| France | 2-5 December 2020 | 1,058 | 93.6 |
| Germany | 5-9 December 2020 | 2,091 | 36.1 |
| Italy | 5-7 December 2020 | 1,025 | 118.5 |
| New Zealand | 5-9 December 2020 | 1,011 | 0.5 |
| Poland | 5-8 December 2020 | 1,023 | 71.3 |
| Sweden | 5-9 December 2020 | 1,016 | 81.3 |
| UK | 5-8 December 2020 | 1,031 | 106.5 |
| USA | 4-11 December 2020 | 2,008 | 101.5 |

Note: Deaths per 100,000 population on December 28th 2020 from https://coronavirus.jhu.edu/data/mortality

**Table S2.** Gender Differences on COVID-19 Vaccination

| | (1) | (2) | (3) | (4) | (5) | (6) | (7) | (8) | (9) |
|---|---|---|---|---|---|---|---|---|---|
| VARIABLES | Vaccination is the only Solution | | | Agree to be Vaccinated | | | Agree to Compulsory Vaccination | | |
| Women | -0.035*** | -0.030*** | -0.033*** | -0.079*** | -0.074*** | -0.076*** | -0.064*** | -0.061*** | -0.067*** |
| | [0.005] | [0.005] | [0.005] | [0.006] | [0.006] | [0.006] | [0.009] | [0.009] | [0.009] |
| No High School | | -0.056*** | -0.026** | | -0.083*** | -0.048*** | | -0.050*** | -0.024 |
| | | [0.011] | [0.011] | | [0.013] | [0.012] | | [0.018] | [0.018] |
| High School | | -0.026*** | -0.006 | | -0.057*** | -0.033*** | | -0.028*** | -0.010 |
| | | [0.006] | [0.006] | | [0.007] | [0.007] | | [0.011] | [0.011] |
| Service Workers | | -0.003 | -0.002 | | -0.024** | -0.022** | | -0.027* | -0.025* |
| | | [0.008] | [0.007] | | [0.009] | [0.009] | | [0.015] | [0.014] |
| Blue Collars | | -0.025*** | -0.014 | | -0.067*** | -0.047*** | | -0.033* | -0.021 |
| | | [0.010] | [0.009] | | [0.012] | [0.011] | | [0.017] | [0.017] |
| Inactive | | 0.001 | 0.004 | | -0.019* | -0.011 | | -0.005 | -0.002 |
| | | [0.009] | [0.008] | | [0.010] | [0.009] | | [0.016] | [0.015] |
| 35-49 yo | | 0.028*** | 0.016** | | 0.002 | -0.009 | | -0.028** | -0.040*** |
| | | [0.008] | [0.007] | | [0.009] | [0.008] | | [0.013] | [0.013] |
| 50-59 yo | | 0.077*** | 0.053*** | | 0.062*** | 0.037*** | | 0.041*** | 0.019 |
| | | [0.009] | [0.008] | | [0.010] | [0.009] | | [0.014] | [0.014] |
| 60+ yo | | 0.133*** | 0.088*** | | 0.134*** | 0.085*** | | 0.069*** | 0.024* |
| | | [0.008] | [0.008] | | [0.009] | [0.009] | | [0.013] | [0.014] |
| Risk Aversion | | | 0.002** | | | -0.003** | | | -0.004** |
| | | | [0.001] | | | [0.001] | | | [0.002] |
| COVID serious health issue | | | 0.060*** | | | 0.038*** | | | 0.074*** |
| | | | [0.006] | | | [0.007] | | | [0.011] |
| Probability of getting infected | | | 0.010*** | | | 0.014*** | | | 0.010*** |
| | | | [0.001] | | | [0.001] | | | [0.002] |
| Probability of serious ill if infected | | | 0.016*** | | | 0.021*** | | | 0.018*** |
| | | | [0.001] | | | [0.001] | | | [0.002] |
| Trust in Scientists | | | 0.184*** | | | 0.208*** | | | 0.149*** |
| | | | [0.008] | | | [0.009] | | | [0.012] |
| Liberal ideology | | | 0.010 | | | 0.049*** | | | 0.066*** |
| | | | [0.007] | | | [0.009] | | | [0.014] |
| Centrist ideology | | | -0.012* | | | 0.011 | | | 0.027** |
| | | | [0.006] | | | [0.007] | | | [0.012] |
| Unknown ideology | | | -0.030*** | | | -0.031** | | | 0.001 |
| | | | [0.011] | | | [0.012] | | | [0.018] |
| Constant | 0.753*** | 0.699*** | 0.368*** | 0.736*** | 0.730*** | 0.238 | 0.490*** | 0.499*** | 0.362 |
| | [0.017] | [0.018] | [0.117] | [0.018] | [0.019] | [0.189] | [0.030] | [0.033] | [0.237] |
| Country-Region FE | yes | yes | yes | yes | yes | yes | yes | yes | yes |
| Observations | 13,019 | 12,914 | 12,498 | 11,918 | 11,838 | 11,628 | 12,126 | 12,041 | 11,831 |
| R-squared | 0.035 | 0.072 | 0.206 | 0.091 | 0.132 | 0.272 | 0.068 | 0.077 | 0.124 |

Notes: Robust standard errors are in parentheses (***, **, * indicate significance at 1, 5, and 10 percent, respectively). We pool survey data from the ten countries in the sample.

**Table S3.** Vaccination is the only Permanent Solution to COVID-19

| | (1) | (2) | (3) | (4) | (5) | (6) | (7) | (8) | (9) | (10) | (11) |
|---|---|---|---|---|---|---|---|---|---|---|---|
| VARIABLES | ALL | AUS | AUT | FRA | GER | ITA | NZ | POL | SW | UK | US |
| Women | -0.030*** | -0.038** | -0.052** | -0.042*** | -0.060*** | 0.021 | -0.018 | -0.041** | -0.018 | 0.013 | -0.031** |
| | [0.005] | [0.018] | [0.022] | [0.013] | [0.013] | [0.018] | [0.018] | [0.020] | [0.018] | [0.018] | [0.014] |
| No High School | -0.056*** | 0.020 | -0.060* | -0.050 | -0.051 | -0.084*** | -0.034 | -0.101 | -0.034 | -0.076** | -0.082* |
| | [0.010] | [0.026] | [0.032] | [0.036] | [0.034] | [0.032] | [0.028] | [0.063] | [0.028] | [0.039] | [0.042] |
| High School | -0.026*** | 0.010 | -0.008 | -0.029** | -0.032** | -0.029 | -0.044** | -0.019 | -0.044** | -0.031 | -0.040*** |
| | [0.006] | [0.022] | [0.034] | [0.014] | [0.015] | [0.021] | [0.020] | [0.023] | [0.020] | [0.020] | [0.015] |
| Service Workers | -0.003 | -0.017 | -0.033 | -0.004 | 0.027 | 0.035 | 0.007 | -0.036 | 0.007 | -0.010 | 0.006 |
| | [0.008] | [0.024] | [0.036] | [0.021] | [0.023] | [0.037] | [0.026] | [0.036] | [0.026] | [0.025] | [0.018] |
| Blue Collars | -0.025*** | -0.074*** | -0.072* | -0.022 | -0.036 | 0.021 | 0.043 | -0.035 | 0.043 | -0.028 | -0.002 |
| | [0.009] | [0.028] | [0.040] | [0.027] | [0.026] | [0.040] | [0.033] | [0.036] | [0.033] | [0.031] | [0.020] |
| Inactive | 0.001 | -0.068 | -0.013 | -0.002 | 0.015 | 0.033 | 0.026 | 0.016 | 0.026 | 0.003 | 0.020 |
| | [0.009] | [0.046] | [0.039] | [0.024] | [0.022] | [0.035] | [0.029] | [0.033] | [0.029] | [0.028] | [0.029] |
| 35-49 yo | 0.028*** | 0.017 | 0.041 | -0.011 | 0.028 | 0.035 | 0.014 | 0.056** | 0.014 | 0.043* | 0.030* |
| | [0.007] | [0.024] | [0.029] | [0.018] | [0.019] | [0.025] | [0.023] | [0.026] | [0.023] | [0.024] | [0.018] |
| 50-59 yo | 0.077*** | 0.007 | 0.106*** | 0.051** | 0.094*** | 0.064** | 0.049* | 0.171*** | 0.049* | 0.082*** | 0.034 |
| | [0.008] | [0.026] | [0.033] | [0.021] | [0.021] | [0.029] | [0.027] | [0.029] | [0.027] | [0.028] | [0.022] |
| 60+ yo | 0.133*** | 0.128*** | 0.109*** | 0.122*** | 0.137*** | 0.103*** | 0.090*** | 0.146*** | 0.090*** | 0.161*** | 0.147*** |
| | [0.007] | [0.026] | [0.033] | [0.020] | [0.019] | [0.025] | [0.025] | [0.027] | [0.025] | [0.026] | [0.017] |
| Constant | 0.699*** | 0.738*** | 0.723*** | 0.687*** | 0.652*** | 0.662*** | 0.700*** | 0.581*** | 0.700*** | 0.715*** | 0.696*** |
| | [0.017] | [0.028] | [0.063] | [0.024] | [0.027] | [0.039] | [0.028] | [0.037] | [0.028] | [0.039] | [0.023] |
| Country-Region FE | Yes | yes | yes | yes | yes | yes | yes | yes | yes | yes | yes |
| Observations | 12,914 | 894 | 994 | 2,082 | 2,091 | 1,025 | 1,011 | 1,023 | 1,011 | 1,016 | 1,747 |
| R-squared | 0.072 | 0.061 | 0.048 | 0.056 | 0.071 | 0.034 | 0.050 | 0.072 | 0.050 | 0.065 | 0.058 |

Notes: Robust standard errors are in parentheses (***, **, * indicate significance at 1, 5, and 10 percent, respectively).

**Table S4**: Agreement to be Vaccinated

| VARIABLES | (1) ALL | (2) AUS | (3) AUT | (4) FRA | (5) GER | (6) ITA | (7) NZ | (8) POL | (9) SW | (10) UK | (11) US |
|---|---|---|---|---|---|---|---|---|---|---|---|
| Women | -0.074*** | -0.062*** | -0.094*** | -0.123*** | -0.076*** | -0.058*** | -0.055*** | -0.088*** | -0.055*** | -0.070*** | -0.059*** |
| | [0.006] | [0.020] | [0.024] | [0.021] | [0.015] | [0.020] | [0.021] | [0.021] | [0.021] | [0.021] | [0.016] |
| No High School | -0.083*** | -0.004 | -0.061* | -0.054 | -0.026 | -0.150*** | -0.092*** | -0.022 | -0.092*** | -0.098** | -0.198*** |
| | [0.012] | [0.029] | [0.035] | [0.060] | [0.040] | [0.035] | [0.033] | [0.068] | [0.033] | [0.044] | [0.046] |
| High School | -0.057*** | -0.024 | -0.026 | -0.081*** | -0.057*** | -0.062*** | -0.056** | -0.013 | -0.056** | -0.052** | -0.084*** |
| | [0.007] | [0.025] | [0.038] | [0.024] | [0.018] | [0.023] | [0.024] | [0.024] | [0.024] | [0.023] | [0.017] |
| Service Workers | -0.024** | -0.032 | -0.079* | -0.019 | -0.015 | 0.016 | -0.003 | -0.032 | -0.003 | -0.087*** | -0.000 |
| | [0.009] | [0.026] | [0.040] | [0.034] | [0.026] | [0.041] | [0.031] | [0.039] | [0.031] | [0.029] | [0.020] |
| Blue Collars | -0.067*** | -0.129*** | -0.126*** | -0.128*** | -0.090*** | 0.015 | 0.030 | -0.051 | 0.030 | -0.084** | -0.047** |
| | [0.011] | [0.031] | [0.044] | [0.045] | [0.030] | [0.044] | [0.039] | [0.039] | [0.039] | [0.035] | [0.023] |
| Inactive | -0.019* | -0.054 | -0.055 | -0.012 | -0.016 | 0.011 | 0.018 | -0.021 | 0.018 | -0.009 | -0.042 |
| | [0.010] | [0.051] | [0.043] | [0.040] | [0.026] | [0.039] | [0.034] | [0.035] | [0.034] | [0.032] | [0.034] |
| 35-49 yo | 0.002 | 0.024 | -0.023 | -0.012 | 0.019 | -0.009 | 0.004 | 0.023 | 0.004 | 0.021 | -0.012 |
| | [0.008] | [0.026] | [0.032] | [0.030] | [0.022] | [0.027] | [0.027] | [0.028] | [0.027] | [0.028] | [0.020] |
| 50-59 yo | 0.062*** | 0.023 | 0.052 | 0.094*** | 0.101*** | 0.027 | 0.008 | 0.110*** | 0.008 | 0.052 | 0.048* |
| | [0.009] | [0.029] | [0.036] | [0.034] | [0.024] | [0.032] | [0.031] | [0.031] | [0.031] | [0.032] | [0.026] |
| 60+ yo | 0.134*** | 0.146*** | 0.102*** | 0.122*** | 0.164*** | 0.087*** | 0.093*** | 0.170*** | 0.093*** | 0.165*** | 0.108*** |
| | [0.008] | [0.029] | [0.037] | [0.032] | [0.022] | [0.028] | [0.029] | [0.029] | [0.029] | [0.030] | [0.020] |
| Constant | 0.730*** | 0.790*** | 0.705*** | 0.557*** | 0.596*** | 0.705*** | 0.702*** | 0.551*** | 0.702*** | 0.768*** | 0.738*** |
| | [0.020] | [0.031] | [0.069] | [0.039] | [0.032] | [0.043] | [0.033] | [0.040] | [0.033] | [0.044] | [0.026] |
| Country-Region FE | yes | yes | yes | | yes | yes | yes | yes | yes | yes | yes |
| Observations | 11,838 | 864 | 994 | 1,042 | 2,091 | 1,025 | 1,011 | 1,023 | 1,011 | 1,016 | 1,741 |
| R-squared | 0.132 | 0.087 | 0.054 | 0.096 | 0.088 | 0.046 | 0.055 | 0.073 | 0.055 | 0.109 | 0.073 |

Notes: Robust standard errors are in parentheses (***, **, * indicate significance at 1, 5, and 10 percent, respectively).

**Table S5**: Agreement with Compulsory Vaccination

| | (1) | (2) | (3) | (4) | (5) | (6) | (7) | (8) | (9) | (10) | (11) |
|---|---|---|---|---|---|---|---|---|---|---|---|
| VARIABLES | ALL | AUS | AUT | FRA | GER | ITA | NZ | POL | SW | UK | US |
| Women | -0.061*** | 0.006 | -0.114*** | -0.127*** | -0.054** | -0.018 | -0.040 | -0.137*** | -0.040 | -0.067** | -0.015 |
| | [0.009] | [0.033] | [0.028] | [0.028] | [0.021] | [0.032] | [0.032] | [0.028] | [0.032] | [0.032] | [0.024] |
| No High School | -0.050*** | -0.002 | -0.073* | -0.047 | 0.040 | -0.188*** | -0.036 | 0.121 | -0.036 | 0.019 | -0.171** |
| | [0.017] | [0.047] | [0.041] | [0.078] | [0.055] | [0.056] | [0.051] | [0.089] | [0.051] | [0.069] | [0.071] |
| High School | -0.028*** | 0.001 | -0.042 | -0.063** | 0.032 | -0.102*** | -0.017 | 0.014 | -0.017 | -0.000 | -0.092*** |
| | [0.010] | [0.041] | [0.044] | [0.031] | [0.024] | [0.038] | [0.037] | [0.032] | [0.037] | [0.035] | [0.026] |
| Service Workers | -0.027* | -0.061 | -0.052 | 0.004 | -0.065* | 0.140** | -0.124*** | -0.035 | -0.124*** | -0.038 | -0.005 |
| | [0.014] | [0.044] | [0.047] | [0.044] | [0.036] | [0.066] | [0.048] | [0.051] | [0.048] | [0.045] | [0.031] |
| Blue Collars | -0.033** | -0.057 | -0.083 | -0.048 | -0.068* | 0.112 | -0.057 | -0.086* | -0.057 | -0.069 | 0.034 |
| | [0.016] | [0.051] | [0.051] | [0.058] | [0.041] | [0.071] | [0.060] | [0.051] | [0.060] | [0.055] | [0.034] |
| Inactive | -0.005 | 0.098 | -0.014 | 0.001 | -0.047 | 0.088 | -0.095* | 0.011 | -0.095* | 0.030 | -0.012 |
| | [0.015] | [0.085] | [0.051] | [0.052] | [0.036] | [0.063] | [0.053] | [0.047] | [0.053] | [0.049] | [0.050] |
| 35-49 yo | -0.028** | 0.057 | -0.003 | 0.027 | -0.016 | -0.074* | -0.067 | 0.020 | -0.067 | -0.052 | -0.088*** |
| | [0.012] | [0.043] | [0.037] | [0.039] | [0.030] | [0.044] | [0.042] | [0.037] | [0.042] | [0.043] | [0.030] |
| 50-59 yo | 0.041*** | 0.083* | 0.029 | 0.068 | 0.079** | -0.008 | 0.067 | 0.105** | 0.067 | 0.044 | -0.023 |
| | [0.014] | [0.048] | [0.042] | [0.044] | [0.033] | [0.052] | [0.049] | [0.041] | [0.049] | [0.050] | [0.038] |
| 60+ yo | 0.069*** | 0.183*** | 0.053 | 0.059 | 0.139*** | -0.042 | 0.114** | 0.133*** | 0.114** | 0.101** | 0.013 |
| | [0.012] | [0.048] | [0.043] | [0.042] | [0.030] | [0.045] | [0.045] | [0.038] | [0.045] | [0.046] | [0.030] |
| Constant | 0.499*** | 0.579*** | 0.266*** | 0.294*** | 0.302*** | 0.527*** | 0.674*** | 0.341*** | 0.674*** | 0.469*** | 0.526*** |
| | [0.028] | [0.051] | [0.081] | [0.051] | [0.044] | [0.070] | [0.052] | [0.053] | [0.052] | [0.069] | [0.040] |
| Country-Region FE | yes | yes | yes | yes | yes | yes | yes | yes | yes | yes | yes |
| Observations | 12,041 | 932 | 992 | 1,042 | 2,089 | 1,024 | 1,010 | 1,021 | 1,010 | 1,016 | 1,884 |
| R-squared | 0.077 | 0.031 | 0.037 | 0.052 | 0.034 | 0.024 | 0.051 | 0.070 | 0.051 | 0.039 | 0.020 |

Notes: Robust standard errors are in parentheses (***, **, * indicate significance at 1, 5, and 10 percent, respectively).

**Table S6**: Gender differences in Attitudinal Factors

| VARIABLES | (1)<br>Risk Aversion | (2)<br>COVID Serious Health Concern | (3)<br>Probability of be Infected | (4)<br>Probability of serious ill if Infected | (5)<br>Trust in Scientists | (6)<br>Liberal Ideology | (7)<br>Central Ideology | (8)<br>Vaccination is the Solution | (9)<br>Multinationals' Fault | (10)<br>Parents of School Kids |
|---|---|---|---|---|---|---|---|---|---|---|
| Women | 0.566*** | 0.052*** | 0.175*** | 0.089* | -0.008 | 0.026*** | -0.005 | -0.030*** | 0.143** | 0.006 |
| | [0.045] | [0.008] | [0.048] | [0.048] | [0.007] | [0.007] | [0.009] | [0.005] | [0.057] | [0.006] |
| No High School | 0.009 | 0.013 | -0.545*** | -0.080 | -0.105*** | -0.083*** | -0.005 | -0.056*** | 1.145*** | 0.035*** |
| | [0.091] | [0.016] | [0.097] | [0.096] | [0.014] | [0.014] | [0.017] | [0.010] | [0.116] | [0.012] |
| High School | 0.115** | -0.000 | -0.396*** | -0.011 | -0.076*** | -0.040*** | 0.008 | -0.026*** | 0.662*** | 0.031*** |
| | [0.052] | [0.009] | [0.055] | [0.055] | [0.008] | [0.008] | [0.010] | [0.006] | [0.065] | [0.007] |
| Service Workers | 0.217*** | -0.002 | -0.120 | 0.003 | 0.001 | 0.004 | 0.002 | -0.003 | 0.150* | -0.021** |
| | [0.070] | [0.012] | [0.074] | [0.074] | [0.010] | [0.011] | [0.013] | [0.008] | [0.088] | [0.010] |
| Blue Collars | 0.175** | 0.007 | -0.385*** | -0.081 | -0.048*** | 0.013 | -0.009 | -0.025*** | 0.491*** | -0.020* |
| | [0.081] | [0.014] | [0.086] | [0.085] | [0.012] | [0.013] | [0.015] | [0.009] | [0.103] | [0.011] |
| Inactive | 0.480*** | 0.024* | -0.345*** | 0.282*** | -0.027** | 0.010 | -0.008 | 0.001 | 0.042 | -0.122*** |
| | [0.076] | [0.013] | [0.081] | [0.081] | [0.011] | [0.012] | [0.015] | [0.009] | [0.096] | [0.010] |
| 35-49 yo | 0.398*** | 0.057*** | 0.059 | 0.478*** | -0.005 | -0.031*** | 0.024** | 0.028*** | 0.032 | 0.181*** |
| | [0.061] | [0.010] | [0.065] | [0.064] | [0.009] | [0.010] | [0.012] | [0.007] | [0.077] | [0.008] |
| 50-59 yo | 0.617*** | 0.085*** | 0.003 | 0.806*** | 0.017 | -0.006 | 0.035** | 0.077*** | -0.558*** | 0.006 |
| | [0.071] | [0.012] | [0.075] | [0.075] | [0.011] | [0.011] | [0.014] | [0.008] | [0.089] | [0.010] |
| 60+ yo | 0.530*** | 0.113*** | -0.009 | 1.498*** | 0.059*** | -0.002 | 0.008 | 0.133*** | -1.080*** | -0.097*** |
| | [0.063] | [0.011] | [0.067] | [0.066] | [0.009] | [0.010] | [0.012] | [0.007] | [0.080] | [0.009] |
| Constant | 4.097*** | 0.411*** | 5.908*** | 5.090*** | 0.865*** | 0.193*** | 0.366*** | 0.699*** | 3.141*** | -0.004 |
| | [0.146] | [0.025] | [0.157] | [0.156] | [0.022] | [0.023] | [0.028] | [0.017] | [0.198] | [0.020] |
| Country-Region FE | yes | yes | yes | yes | yes | yes | yes | yes | yes | yes |
| Observations | 12,808 | 13,210 | 13,215 | 13,215 | 13,210 | 13,215 | 13,215 | 12,914 | 12,763 | 13,215 |
| R-squared | 0.083 | 0.091 | 0.089 | 0.073 | 0.049 | 0.020 | 0.036 | 0.072 | 0.096 | 0.191 |

Notes: Robust standard errors are in parentheses (***, **, * indicate significance at 1, 5, and 10 percent, respectively). We pool survey data from the ten countries in the sample.

**Table S7**: Gender Differences in Vaccination – Skepticism in COVID-19 Vaccines

| VARIABLES | (1) be Vaccinated | (2) Agree to Compulsory Vaccination |
|---|---|---|
| Women | -0.052*** | -0.050*** |
| | [0.005] | [0.009] |
| Vaccine Solution | 0.534*** | 0.351*** |
| | [0.011] | [0.016] |
| COVID Multinat's Fault | -0.016*** | -0.009*** |
| | [0.001] | [0.001] |
| Schoolkids Parents | 0.012* | 0.008 |
| | [0.007] | [0.012] |
| Constant | 0.144 | 0.290 |
| | [0.152] | [0.216] |
| Country-Region FE | yes | yes |
| Socio-demo controls | yes | yes |
| Attitudinal controls | yes | yes |
| Observations | 11,272 | 11,291 |
| R-squared | 0.473 | 0.170 |

Notes: Robust standard errors are in parentheses (***, **, * indicate significance at 1, 5, and 10 percent, respectively). We pool survey data from the ten countries in the sample. We control for country-region fixed effects and sociodemographic characteristics in all regressions. The sociodemographic controls include age groups (18-34, 35-49, 50-59, 60+), education (no high school, high school, college), occupation status (blue collar, service, white collar, no occupation). Attitudinal controls include risk aversion, probability of being infected, probability of being seriously ill if infected, seriousness of COVID-19 as health risk, trust in scientists and political ideology (left, center, no ideology).
